# TGFβ1 plasma levels and clot lysis assay to better characterize patients after a first unprovoked episode of pulmonary embolism

**DOI:** 10.1101/2024.05.15.24306886

**Authors:** Marc Danguy des Deserts, Claire de Moreuil, Jamal Elhasnaoui, Lenaïck Gourhant, Virginie Gourdou-Latyszenok, Benjamin Espinasse, Juliette Menguy, Cécile Tromeur, Rozenn Le Corre, Raphael Le Mao, Daniel Kraemmer, Olivier Sanchez, PADIS-PE investigators, Francis Couturaud, Catherine A. Lemarié

**Author notes:** **Corresponding author** Catherine A Lemarié, Brest medical faculty, 22, rue Camille Desmoulins, 29609 Brest, France.

## Abstract

**Introduction:** The pathophysiology of residual pulmonary vascular obstruction (RPVO) and recurrent venous thromboembolism (VTE) after unprovoked pulmonary embolism (PE) remains poorly understood. The purpose was to evaluate fibrinolytic and tissue remodeling markers as indicators of RPVO and recurrence after a first unprovoked PE.

**Methods:** Analyses were conducted in the 18 to 70-year-old patients included in the PADIS-PE trial, with a pulmonary vascular obstruction (PVO) index ≥30% at PE diagnosis. After an initial six-month vitamin K antagonist treatment, patients were randomised to receive placebo or warfarin for 18 months, assessed for the absence or presence of residual pulmonary vascular obstruction (RPVO < or ≥5%, respectively) and followed during two years. Quantitative assessment of fibrinolytic (D-dimer, tPA, uPA, TFPI) and tissue remodeling (TGFß1) markers, and a tissue-factor-based turbidimetric clot lysis assay (CLA) were performed one month after warfarin discontinuation.

**Results:** Among the 371 patients included in PADIS-PE, 23 were eligible. Six (26%) patients presented RPVO ≥ 5%, symptomatic recurrent VTE occurred in nine (39%) patients. Clot formation and lysis parameters were not associated with RPVO. TGFß1 plasma levels were higher in patients with RPVO. Clot formation potential measured with CLA was higher in patients with recurrent VTE. No association between recurrent VTE and TGFß1 was observed. In multivariable analysis, time to peak was associated with VTE recurrence.

**Conclusion:** In adult patients with a first unprovoked PE and a PVO index ≥30%, TGFß1 plasma level was associated with RPVO, whereas clot formation parameters measured with CLA were associated with VTE recurrence.

## Introduction

Pulmonary embolism (PE), the most severe clinical presentation of venous thromboembolism (VTE), occurs in one patient over 1000 every year and represents the third leading cause of cardiovascular death [1–3]. The main complications of PE are recurrent non-fatal and fatal VTE, and long-term sequelae (chronic thromboembolic pulmonary disease with or without pulmonary hypertension) [4]. In 50% of cases, PE occurs in the absence of clinical circumstances (termed “unprovoked PE”) [5]. In these patients, the risk of recurrence is high (10% at one year and 35% at 10 years after anticoagulation discontinuation) and guidelines recommend, with a moderate grade of evidence, an indefinite duration of anticoagulation in the absence of a high risk of bleeding [6]. However, if indefinite anticoagulation is highly effective, it exposes patients who would not have experienced recurrent VTE after stopping treatment to an unnecessary continuous increase in the risk of bleeding [6–8]. Therefore, identification of risk factors for VTE recurrence is crucial to avoid long-term anticoagulation in patients at low risk of recurrence.

The development of chronic thromboembolic pulmonary disease without pulmonary hypertension, which is diagnosed by the presence of residual pulmonary vascular obstruction (RPVO) on ventilation/perfusion (V/Q) lung scan [6], occurs in 25% to 66% of patients after at least three months of full therapeutic dose of anticoagulant [9]. Pulmonary vascular obstruction (PVO) index at PE diagnosis and unprovoked PE are associated with RPVO [10]. However, the mechanisms that contribute to the development of RPVO and/or promote recurrent VTE are unknown. An impaired balance between coagulation and fibrinolysis may contribute to RPVO and recurrent VTE [11–14]. Elevated plasma factor VIII, a procoagulant factor, is associated with recurrent VTE [14] and RPVO after PE [15,16]. Decreased plasma fibrinolytic potential, as assessed by prolonged clot lysis time (CLT) measured during clot lysis assay (CLA) [17], is variably associated with recurrent VTE [18–20] and RPVO [21,22] after a first VTE event. In addition, venous thrombus resolution involves a complex interplay between endothelial cells, immune cells and platelets [23]. Excessive activation of endothelial cells, immune cells and platelets can lead to thrombo-inflammation, systemic prothrombotic state and impaired thrombus resolution [24]. Markers of activated endothelium, immune cell activation and persistent thrombi are observed in pulmonary endarterectomy specimens from patients with chronic thromboembolic pulmonary hypertension (CTEPH) [25]. Importantly, CTEPH has been associated with transforming growth factor beta 1 (TGFß1)-dependent mechanisms [26]. However, the association between clot lysis time, recurrent VTE, TGFß1 and RVPO in chronic thromboembolic pulmonary disease without hypertension has not yet been assessed.

The objectives of this exploratory study were to evaluate the association of fibrinolytic parameters and TGFß1 with RPVO and/or with VTE recurrence after a first episode of unprovoked PE following warfarin discontinuation.

## Methods

### Study design of the PADIS-PE trial

This is an ancillary study of the double-blind randomized PADIS-PE trial using the plasma collected for the trial [7]. After written informed consent, patients aged 18 years and older with a first episode of a proven symptomatic unprovoked PE, initially treated during six uninterrupted months with vitamin K antagonists (VKA) were included and randomised to receive either warfarin or placebo for an additional 18 months [7]. All patients were followed for a period of 24 months after study treatment discontinuation [7]. PE was defined as unprovoked if it occurred in the absence of a major reversible risk factor for VTE within three months before PE diagnosis (i.e. surgery with local or general anaesthesia >30 min, trauma with or without lower limb cast, bed rest >72 h) and without active cancer or cancer that had resolved within the last two years before PE diagnosis [7].

At the time of inclusion in the PADIS-PE trial (i.e., after the initial six months of anticoagulation), baseline characteristics of patients were collected and all patients underwent V/Q lung scan acquisition [7]. RPVO was defined as a pulmonary vascular obstruction (PVO) index ≥5% at inclusion (i.e., after six months of anticoagulation) based on V/Q lung scan. All V/Q lung scans were centrally re-interpreted after the last patient achieved complete follow-up of 42 months by two independent readers, who were blinded to study treatment allocation, results of other imaging tests and patient characteristics. Blood was collected from all fasted patients between 8 and 10 am at enrolment and at 1, 18, 19 and 42 months after inclusion. Blood was immediately centrifuged at 2500 g for 15 minutes to obtain platelet poor plasma and stored at – 80° C until use.

The key secondary outcome of the PADIS-PE trial was symptomatic recurrent VTE, including proximal deep vein thrombosis (DVT) and/or PE. All outcomes were adjudicated blindly by an independent central Clinical Events Committee. Patients were enrolled from July 13, 2007 to March 15, 2012 in 14 French hospitals. The PADIS-PE trial was registered on ClinicalTrials.gov under the identifier NCT00740883, the protocol and amendments were approved by a central independent ethics committee [7].

### Patients selection

For this ancillary study, we selected patients enrolled in the PADIS-PE trial: (i) aged from 18 to 70 years; (ii) without major thrombophilia (no protein C, S or antithrombin deficiency, no combined thrombophilia, no antiphospholipid antibodies); (iii) with a PVO index ≥30% at PE diagnosis, based on computed tomography pulmonary angiography or V/Q lung scan; (iiii) and who did not develop CTEPH during the 42-month follow-up period [27]. Patients for whom no warfarin-free blood samples were not available or of poor-quality, and patients for whom PVO index at PE diagnosis and at inclusion (i.e., at 6-month anticoagulation) was missing were excluded from this analysis.

### Plasma clot formation and lysis assay

All blood tests were performed from frozen plasma collected one month and 19 months after enrolment in PADIS-PE in the placebo and warfarin groups respectively (i.e. one month after stopping anticoagulation), to avoid the influence of anticoagulation. CLA was performed following a detailed protocol published previously [28]. Briefly, frozen plasma was thawed in a 37°C water bath for 5 minutes, then 75 µL of plasma were placed in a 96-well plate (NUNC Immunoplate, ThermoFischer Scientific, Cat #10547781). A clot induction and lysis mix was prepared in HEPES-BSA buffer solution (NaCl 150 mM, pH 7,4, HEPES 20 mM, BSA 1%) containing tissue factor (Innovin®, Siemens Health Care Diagnostics, Cat #4212-40), phospholipids (Phospholipid-TGT Reagent, Rossix, n°5-PL604T), CaCl_2_ (1M solution) and human tissue plasminogen activator (tPA, Merck, Cat #T0831). Addition of 75 µL of the clot induction and lysis mix to the plasma and measurement of plasma turbidity (OD 405 nm) were performed using a spectrophotometer (Thermoscientific Varioskan® Flash) at 37°C. Final concentrations of tissue factor, phospholipids, CaCl_2_, and tPA were 1.4 pM, 4 μM, 20 mM, and 333 ng.mL^-1^, respectively. Measurements were performed every 20 seconds for 120 minutes, at 405 nm wavelength with a settle delay of 50 ms.

CLA parameters, as described by Posch *et al.* [28], were obtained from the turbidity curve **(Supplementary Figure 1)**. Clot formation parameters included baseline absorbance, lagphase, maximum clot formation rate (max CFR), time to max CFR, peak absorbance, time to peak and amplitude. Clot lysis parameters comprised CLT, time to 50% clot lysis (t50% lysis), maximum lysis rate (max lysis rate) and time to max lysis rate. Baseline was set as the first optical density (OD) measurement, CLT was set as time from 50% clotting to 50% lysis absorbance. Calculation of these parameters was performed from raw OD measurements using a custom R script, as described by Posch *et al.* [28].

For reproducibility reasons, a validation study included 10 separate CLA runs with 12 replicates of healthy pooled plasma (for details see Supplementary Materials and Methods – Section 1). The intra and inter-assay coefficients of variation of CLA parameters ranged from 2% for the baseline OD to 16% for t50% lysis **(Supplementary Table 1)**.

### Quantitative assessment of fibrinolysis and TGFß1

Tissue factor pathway inhibitor (TFPI), tPA and urokinase-like plasminogen activator (uPA) antigens were measured using ELISA kits (human TFPI, Sigma-Aldrich, Cat #RAB0552, ZYMUTEST tPA antigen, Hyphen Biomed, Cat #RK011A and human PLAU, Sigma-Aldrich, Cat #RAB0555, respectively) according to the manufacturer’s instructions. Samples were analysed using a spectrophotometer (Thermo Scientific Multiskan FC®). All D-dimer levels were measured using high sensitivity VIDAS D-dimer test (BioMérieux).

We used the Luminex microbeads array system to quantify circulating levels of TGFß1 [29]. Analyses were carried out according to the manufacturer’s instructions (Millipore) and quantified in Luminex FLEXMAP using the xPONENT software.

### Statistical analysis

Continuous variables were summarised as medians (interquartile range [IQR]), categorical variables were reported as absolute frequencies (percentages). Univariable analyses were performed to select preliminary predictive variables for the multivariable model and to evaluate the association between each potential marker and RPVO or VTE recurrence. Proportions and *p*-values were calculated for each variable. Wilcoxon-Mann-Whitney and χ^2^ test were used for comparison of continuous and categorical variables, respectively.

To investigate the correlation between the different biomarkers measured, a principal component analysis (PCA) was performed. PCA is a statistical method used to decompose a group of correlated variables into a set of uncorrelated variables called principal components. These principal components are then arranged in decreasing order of variance [30]. As our data had missing values (four for TGFß1 and three for TFPI) **(Supplementary Figure 2)** we imputed a complete dataset prior to PCA using the missMDA R package [31,32]. Briefly, the package allows the use of principal component methods for an incomplete dataset. The missing values are predicted using the iterative PCA algorithm for a predefined number of dimensions. The estim_ncpPCA() function of the missMDA package was used to estimate the number of needed dimensions to impute the number of principal components from the original incomplete dataset. Then, the imputePCA was used to impute a complete dataset using the PCA algorithm while specifying the number of dimensions with the ncp argument. In addition, data were standardized to scale all variables. The PCA() function of the FactoMineR package [31,32] was employed for PCA visualization, and data extraction was performed using corresponding functions from the factoextra package [31,32].

We estimated hazard ratios (HR) (95% confidence intervals (CI)) for VTE recurrence and for RPVO with a multivariable Cox regression model, including variables with a *p*-value <0.2 in univariable analysis.

A two-sided *p*-value <0.05 was considered statistically significant.

Statistical analyses were performed using the R v4.3.3 software (R Foundation for Statistical Computing, Vienna, Austria) or the IBM SPSS Statistics v29.0 software (IBM Corp., Armonk, NY, USA).

## Results

Among the 371 patients enrolled in the PADIS-PE study, 49 were eligible to this ancillary study. Twenty-six patients without warfarin-free samples were excluded. Therefore, 23 (6%) patients were included in the analysis **(Figure 1)**.

**Figure 1.**
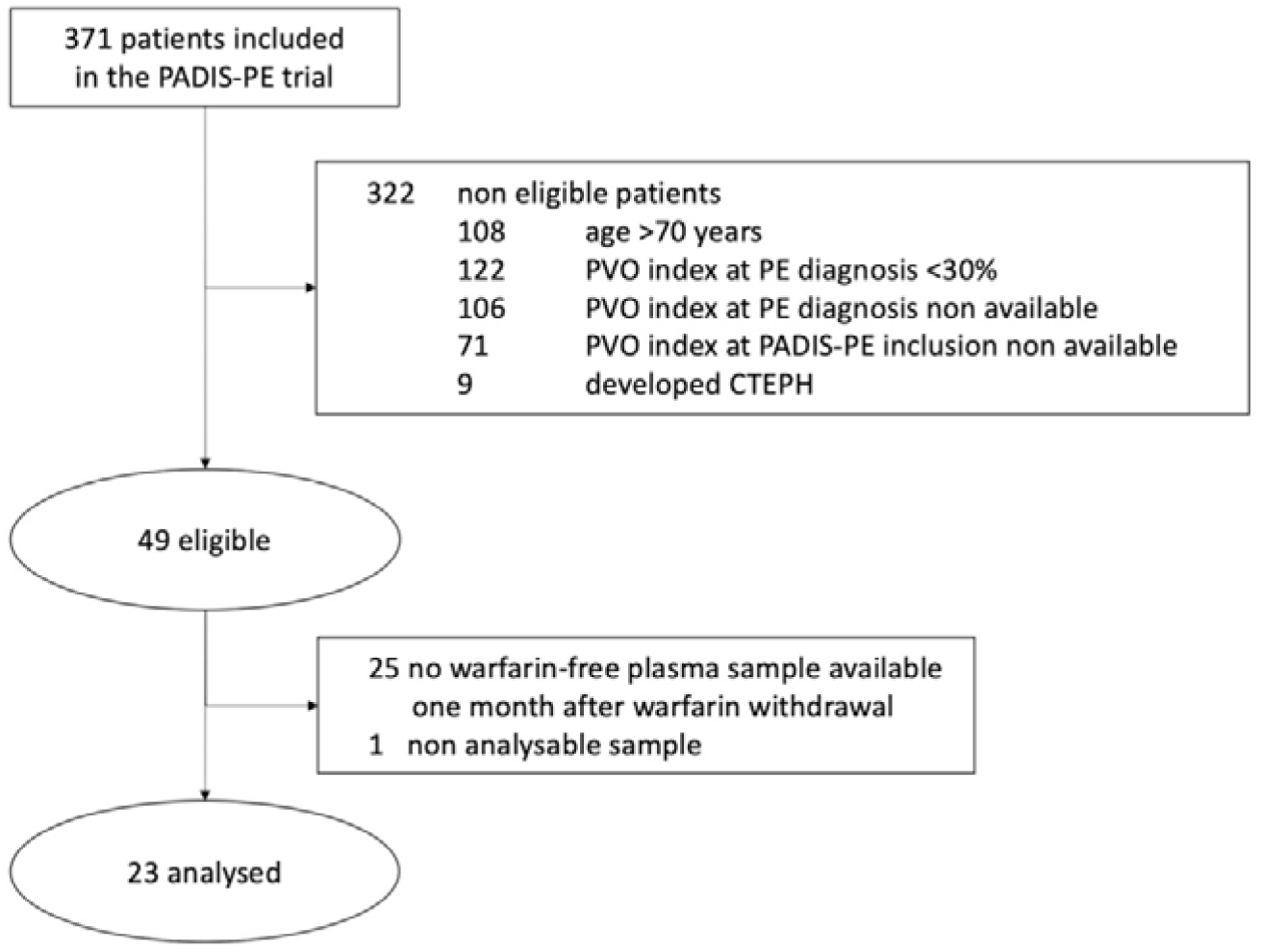
Study flow chart. CTEPH: chronic thromboembolic pulmonary hypertension; PE: pulmonary embolism; PVO: pulmonary vascular obstruction.

### Characteristics of the overall study population

As shown in **Table 1**, median (IQR) age was 56.0 (50.0-59.0) years and seven (30%) patients were female. Median PVO index at PE diagnosis was 57.5% (48.8-62.8). Median follow-up duration was 41 months (41-42). Symptomatic recurrent VTE occurred during follow-up in nine (39%) patients. All events occurred in the absence of anticoagulation, five events being non-fatal PE and four events being isolated DVT.

**Table 1.** Characteristics of the overall population study.

|  | Overall (N=23) |
| --- | --- |
| Age, y | 56.0 (50.0-59.0) |
| Female gender | 7 (30) |
| Body mass index, kg.m<sup>-2</sup> | 27.8 (24.6-30.8) |
| Creatinine clearance, mL.min<sup>-1</sup># | 111.5 (85.9-155.1) |
| Allocated treatment at inclusion |  |
| Placebo | 16 (70) |
| Warfarin | 7 (30) |
| Characteristics of pulmonary embolism |  |
| PVO index at PE diagnosis | 57.5 (48.8-62.8) |
| PVO index ≥5% at PADIS-PE inclusion | 6 (26) |
| VTE recurrence | 9 (39) |
| DVT | 4 (17) |
| PE | 5 (22) |
| Follow-up duration, months | 41 (41-42) |
| Biological assessment one month after warfarin withdrawal |  |
| Clot lysis assay parameters |  |
| Baseline, OD | 0.44 (0.36-0.54) |
| Lagphase, sec | 257 (204-282) |
| Max CFR, OD.min<sup>-1</sup> | 0.29 (0.25-0.40) |
| Time to max CFR, sec | 260 (200-325) |
| Peak absorbance, OD | 1.12 (1.02-1.23) |
| Time to peak absorbance, sec | 490 (440-580) |
| Amplitude, OD | 0.70 (0.59-0.74) |
| Clot lysis time, min | 10.7 (10.1-12.3) |
| Time to 50% clot lysis, min | 8.0 (7.1-9.6) |
| Max lysis rate, OD.min<sup>-1</sup> | 0.06 (0.05-0.08) |
| Time to max lysis rate, min | 15.0 (13.0-16.8) |
| Fibrinolysis markers |  |
| D-dimer, ng.mL<sup>-1</sup> | 480.0 (340.0-832.0) |
| TFPI, ng.mL<sup>-1</sup> | 50.2 (38.0-59.1) |
| tPA, ng.mL<sup>-1</sup> | 5.8 (4.4-7.0) |
| uPA, pg.mL<sup>-1</sup> | 1047.0 (818.3-1337.5) |
| Tissue remodeling marker |  |
| TGFβ1, pg.mL<sup>-1</sup> | 89.1 (64.6-132.2) |

Continuous variables are summarized as medians (interquartile range: IQR), categorical variables are reported as absolute frequencies (percentages). Abbreviations: CFR, clot formation rate; DVT, deep vein thrombosis; OD, optical density; PE, pulmonary embolism; PVO, pulmonary vascular obstruction; TGFβ1, transforming growth factor beta 1; TFPI, tissue factor pathway inhibitor; tPA, tissue-type plasminogen activator ; uPA, urokinase-type plasminogen activator; VTE, venous thromboembolic disease. #: creatinine clearance was calculated using the Cockcroft-Gault formula. †: an anticardiolipin antibody was considered present if either the IgG or IgM antibody titer was more the 99th percentile.

### TGFß1 plasma levels are associated with RPVO

In univariable analysis, there was no association between age, creatinine clearance, sex, body mass index, PVO index at PE diagnosis and the presence of RPVO **(Table 2)**.

**Table 2.**
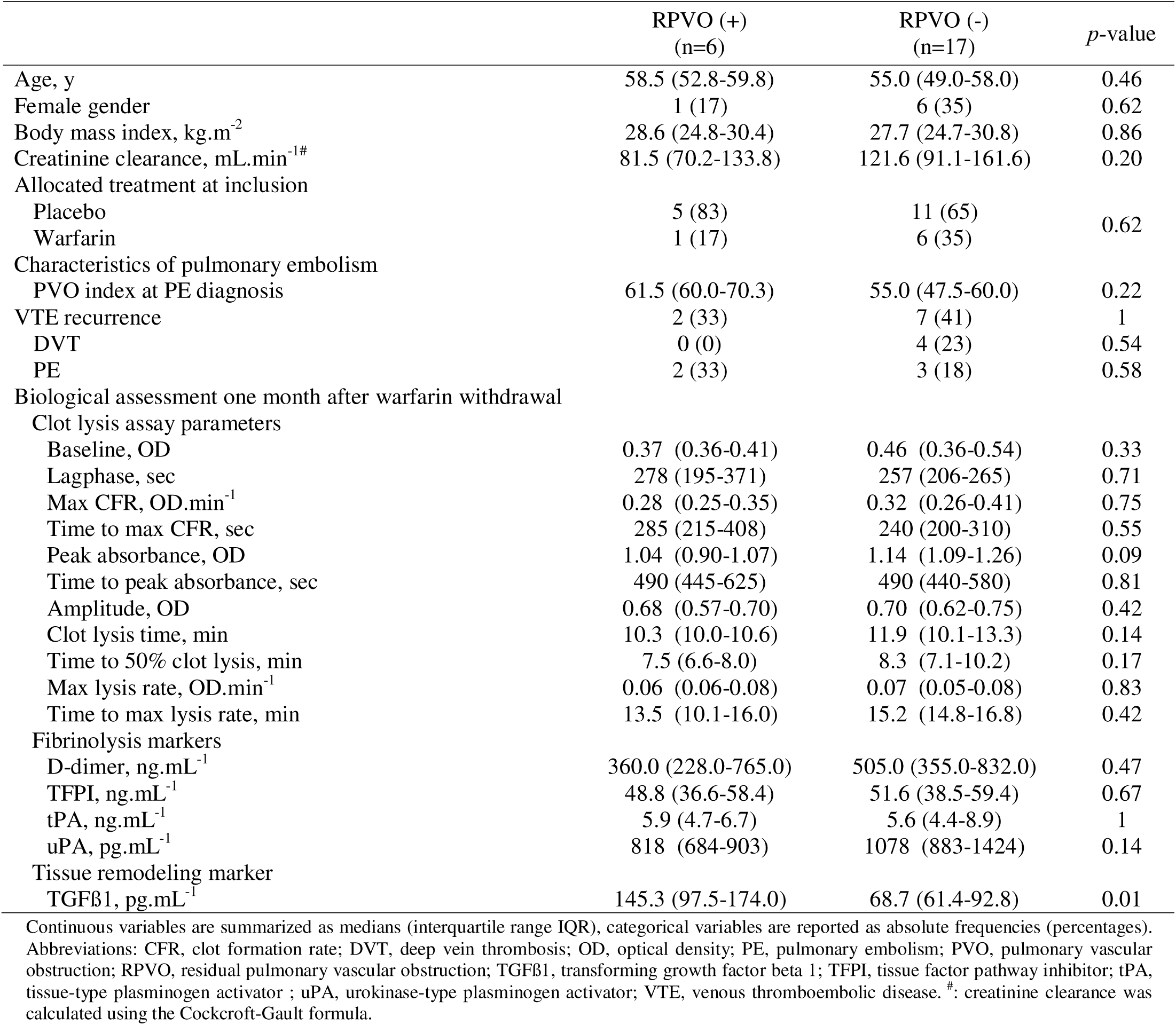
Correlation with residual pulmonary vascular obstruction (RPVO) in univariate analysis.

Neither clot formation nor clot lysis parameters measured by the CLA was associated with the presence of RPVO. Fibrinolysis markers (D-dimer, TFPI, tPA and uPA) were not associated with the presence of RPVO either **(Table 2)**.

Regarding indicator of tissue remodeling, patients presenting RPVO had significantly higher median (IQR) TGFß1 plasma levels (145.3 pg.mL^-1^ [97.5-174.0]) than patients without RPVO (68.7 pg.mL^-1^ [61.4-92.8], *p*=0.01) **(Table 2)**. In the recurrent VTE group, the percentage of RPVO was positively correlated with TGFß1 plasma levels (R^2^=0.56, *p*=0.02) **(Figure 2)**. In the non-recurrent VTE group, there was no correlation between RVPO percentage and TGFß1 plasma levels (R^2^=0.12, *p*=0.34) **(Figure 2)**. Moreover, patients with RVPO and recurrent VTE had the highest median TGFß1 plasma levels compared to patients without RPVO and recurrent VTE (*p*=0.04) **(Supplementary Table 2)**.

**Figure 2.**
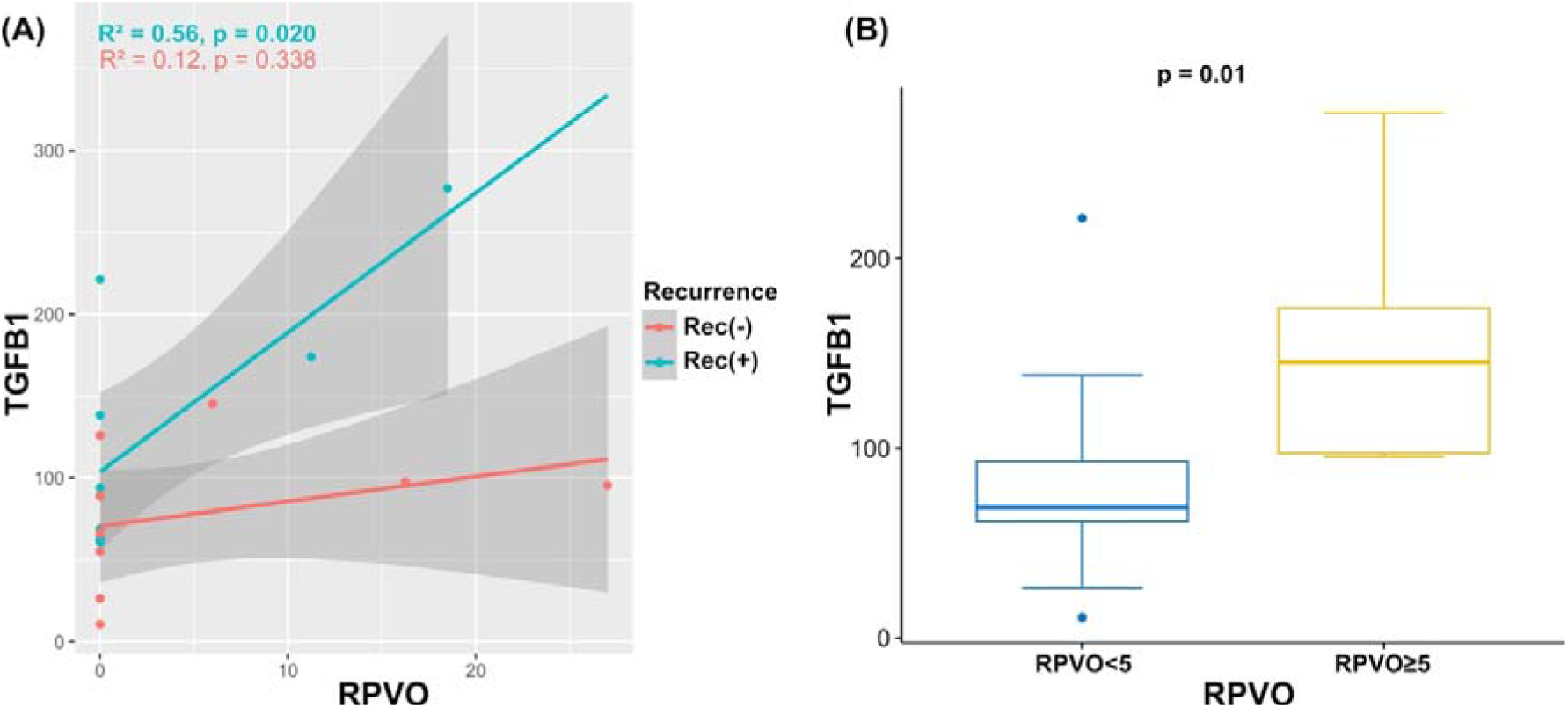
Relationship between TFGB and RPVO. (A) scatterplots showing the degree of correlation between the TGFB1 plasma levels and the quantitatively measured level of residual pulmonary vascular obstruction (RPVO). Each dot represents a patient, with the colour code indicating the recurrence status. (B) TGFB1 plasma levels in patients stratified by the level of residual pulmonary vascular obstruction (e.g. RPVO < 5% and RPVO ≥ 5%). TGFB1: transforming growth factor beta 1; Rec(-): Non-recurrent patient, Rec(+): Recurrent patient.

### CLA parameters are associated with VTE recurrence

In univariable analysis, age and creatinine clearance were found to be associated with VTE recurrence. We found no evidence for a difference regarding sex, body mass index, PVO index at PE diagnosis or at inclusion in the PADIS-PE trial on the risk of VTE recurrence **(Table 3)**.

**Table 3.** Risk factors of recurrent venous thromboembolism in univariate analysis.

|  | Recurrence (+)<br>(n=9) | Recurrence (-)<br>(n=14) | *p*-value |
| --- | --- | --- | --- |
| Age, y | 60.0 (58.0-63.0) | 52.0 (47.5-56.0) | 0.02 |
| Female gender | 2 (22) | 5 (36) | 0.66 |
| Body mass index, $\text{kg.m}^{-2}$ | 25.8 (24.5-29.4) | 27.9 (25.4-30.8) | 0.69 |
| Creatinine clearance, $\text{mL.min}^{-1}\#$ | 88.2 (73.8-104.8) | 144.9 (98.8-169.8) | 0.03 |
| Allocated treatment at inclusion |  |  |  |
| Placebo | 6 (67) | 10 (71) | 1 |
| Warfarin | 3 (33) | 4 (29) |  |
| Characteristics of pulmonary embolism |  |  |  |
| PVO index at PE diagnosis | 57.5 (50.0-75.0) | 57.5 (48.1-60.0) | 0.53 |
| PVO index $\geq 5\%$ at PADIS-PE inclusion | 2 (22) | 4 (29) | 1 |
| Biological assessment one month after warfarin withdrawal |  |  |  |
| Clot lysis assay parameters |  |  |  |
| Baseline, OD | 0.44 (0.37-0.54) | 0.42 (0.33-0.53) | 0.55 |
| Lagphase, sec | 206 (188-243) | 265 (253-334) | 0.01 |
| Max CFR, $\text{OD.min}^{-1}$ | 0.32 (0.27-0.44) | 0.29 (0.24-0.36) | 0.23 |
| Time to max CFR, sec | 200 (180-240) | 310 (245-340) | 0.003 |
| Peak absorbance, OD | 1.14 (1.00-1.27) | 1.12 (1.03-1.19) | 0.68 |
| Time to peak absorbance, sec | 440 (430-460) | 530 (493-618) | 0.007 |
| Amplitude, OD | 0.64 (0.45-0.73) | 0.70 (0.63-0.74) | 0.38 |
| Clot lysis time, min | 11.9 (10.2-13.9) | 10.3 (9.9-12.2) | 0.14 |
| Time to 50% clot lysis, min | 9.1 (7.5-10.4) | 7.4 (6.9-8.9) | 0.10 |
| Max lysis rate, $\text{OD.min}^{-1}$ | 0.05 (0.04-0.06) | 0.07 (0.06-0.08) | 0.01 |
| Time to max lysis rate, min | 15 (9.2-16.5) | 15.1 (13.3-16.8) | 0.73 |
| Fibrinolysis markers |  |  | 0.56 |
| D-dimer, $\text{ng.mL}^{-1}$ | 463.0 (355.0-765.0) | 516.0 (340.0-898.0) | 0.56 |
| TFPI, $\text{ng.mL}^{-1}$ | 46.8 (36.6-58.7) | 51.6 (41.1-59.3) | 0.77 |
| tPA, $\text{ng.mL}^{-1}$ | 5.8 (4.5-8.9) | 5.4 (4.4-7.0) | 0.85 |
| uPA, $\text{pg.mL}^{-1}$ | 857 (747-1145) | 1069 (894-1636) | 0.20 |
| Tissue remodeling marker |  |  |  |
| TGFβ1, $\text{pg.mL}^{-1}$ | 94.1 (68.8-168.2) | 78.9 (57.8-97.0) | 0.21 |

Continuous variables are summarized as medians (interquartile range IQR), categorical variables are reported as absolute frequencies (percentages). Abbreviations: CFR, clot formation rate; DVT, deep vein thrombosis; OD, optical density; PE, pulmonary embolism; PVO, pulmonary vascular obstruction; TGFβ1, transforming growth factor beta 1; TFPI, tissue factor pathway inhibitor; tPA, tissue-type plasminogen activator ; uPA, urokinase-type plasminogen activator; VTE, venous thromboembolic disease. #: creatinine clearance was calculated using the Cockcroft-Gault formula.

Regarding CLA parameters, three clot formation parameters (lagphase, time to max CFR, time to peak) and one clot lysis parameters (max lysis rate) were associated with VTE recurrence. Median (IQR) lagphase, time to max CFR and time to peak were shorter in the recurrent group compared to the non-recurrent group (206 sec [188-243] vs 265 sec [253-334], *p*=0.01; 200 sec [180-240] vs 310 sec [245-340], *p*=0.003; 440 sec [430-460] vs 530 sec [493-618], *p*=0.007, respectively). Median (IQR) max lysis rate was lower in the recurrent group in comparison with the non-recurrent group (0.05 OD.min^-1^ [0.04-0.06] vs 0.07 OD.min^-1^ [0.06-0.08], *p*=0.01). No association was found between baseline absorbance, max CFR, peak absorbance, amplitude, CLT, time to 50% clot lysis, or time to max lysis rate and VTE recurrence **(Table 3)**.

Plasma levels of fibrinolysis biomarkers (D-dimer, TFPI, tPA and uPA) and TGFß1 levels were not associated with VTE recurrence **(Table 3)**.

### Principal component analysis (PCA)

A principal component analysis (PCA) was performed on the 16 measured parameters, including CLA and tissue remodeling biomarkers. As shown in **Figure 3A, B** and **Supplementary Table 3**, a cumulative 83.3% of the variation of these 16 variables was explained by their first five principal components (PCs). PCA was performed considering only these five PCs, since their eigenvalue > 1, meaning they account for more variance than accounted by one of the original variables. The individual contribution of each of the 16 variables to these PCs is reported in **Figure 3C**. Furthermore, 47.6% of the variance was explained by their first two PCs. A variable correlation plot is reported in **Figure 3D** displaying how much the 16 variables are correlated with these components. The quality of representation of each variable on the factorial plane is represented by the length of the arrows. The angle between the variables (their arrows) provides an indication on how well the variables are correlated on the factorial plane (i.e. 90° no correlation, 180° negative correlation). PC1 was mainly composed of clot formation parameters (time to peak absorbance, time to maxCFR, lagphase, max CFR) and one clot lysis parameter (time to max lysis rate). PC2 was mainly composed of clot lysis parameters (CLT, t50% lysis, max lysis rate), fibrinolysis markers (tPA, TFPI) and TGFß1 **(Figure 3C, D)**. Of note, TGFß1 plasma levels and maximum lysis rate were inversely correlated on this factorial map, but whether this inverse correlation has a functional impact on the fibrinolytic system has to be investigated. Other coagulation (baseline OD, peak absorbance, amplitude) and fibrinolysis parameters (D-dimer and uPA) had minor contributions to PC1 and PC2. The individual contributions of each variable to PC1, PC2, or both are reported in **Supplementary Figure 3**.

**Figure 3.**
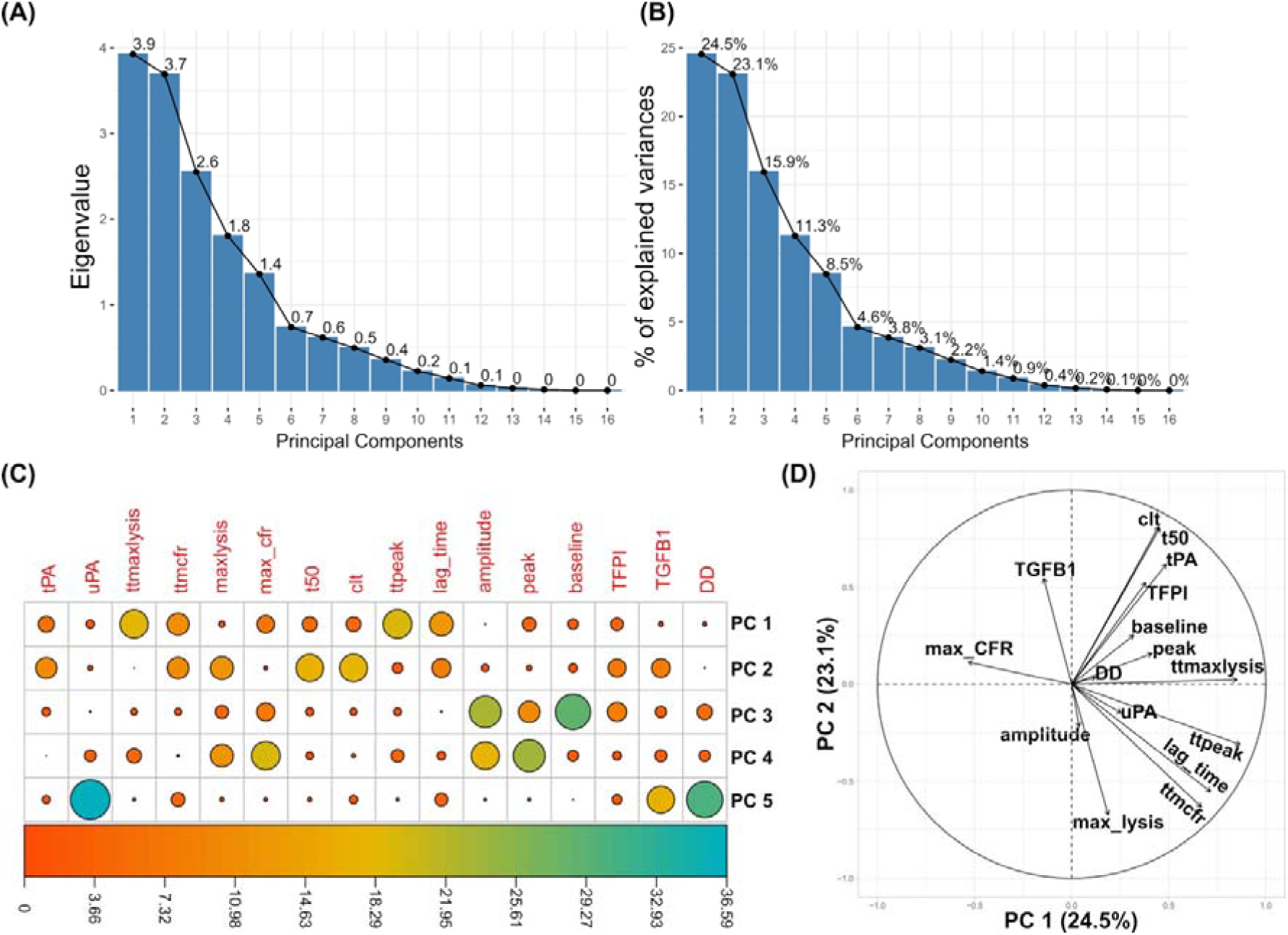
Integrative analysis of the fibrinolysis and CLA parameters by principal component analysis. **(A)** Barplots indicating the eigenvalues of each principal component. The eigenvalues measure the amount of variation retained by each principal component. (**B)** A scree plot with the percentage of explained variance by each principal component. A cumulative proportion of explained variance of 83.3% is obtained at the fifth principal component. **(C)** A correlation plot showing the contribution of each of the 16 measured parameters (in red) to each of the five retained principal components. **(D)** A variable correlation plot showing the projections of the observations as well as the relationships between all 16 variables, with positively correlated plots are grouped together (e.g. clt, t50, tPA, TFPI, etc or TGB1 and max_CFR) and negatively correlated parameters are positioned on opposite sides of the plot origin. Parameters that are distant from the plot origin are well presented by the factor map.

Figure 4 **A and B** display scatter plots of the first two PCs, PC1 as the x-axis and PC2 as the y-axis, stratified by RPVO (Figure 4A) and VTE recurrence (Figure 4B). There was no apparent clustering of patients when stratified by RPVO (Figure 4A), while patients with VTE recurrence had higher values in their second PC on the factorial plane (Figure 4B).

**Figure 4.**
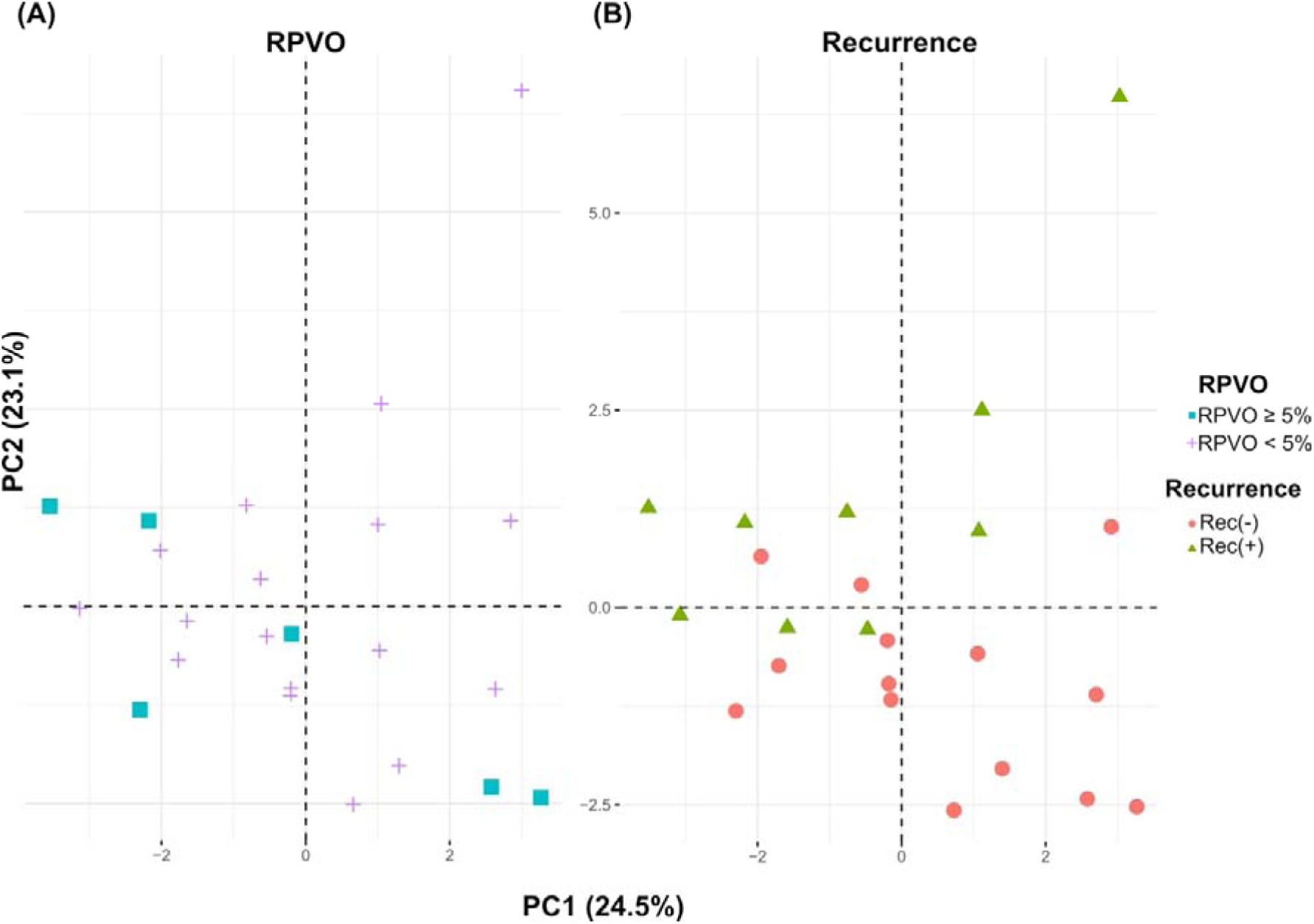
Scatter plots of the first two PCs stratified on (A) the level (%) of residual pulmonary vascular obstruction (RPVO < 5%; RPVO ≥ 5%), and (B) recurrence status. On both (A) and (B), each symbol represents a patient and information on the level of RPVO and recurrence status is given for the same patient, a square or a cross on (A) (patient presenting RPVO <5% or RPVO ≥5%, respectively) and a dot or a triangle on (B) (no recurrent VTE or recurrent VTE, respectively). Abbreviations: PC1, principal component number 1; PC2, principal component number 2; Rec, recurrence; RPVO, residual pulmonary vascular obstruction.

### Multivariable analysis

Seven variables were selected from the univariable analysis (age, time to maxCFR, time to peak absorbance, CLT, t50% lysis, max lysis rate and uPA) to determine the association between each of them and VTE recurrence in the multivariable analysis model. Time to peak was the only parameter independently associated with VTE recurrence (HR 0.97; 95% CI [0.94-0.93], *p=*0.01).

Finally, six variables were selected from the univariable analysis (creatinine clearance, peak absorbance, CLT, t50% lysis, uPA and TGFß1) to determine the association between each of them and the presence of RPVO in the multivariable analysis model. There was no parameter independently associated with the presence of RPVO.

## Discussion

In this ancillary analysis based on 23 unprovoked PE patients enrolled in the PADIS-PE study with a strong thrombotic phenotype (aged from 18 to 70 years, PVO index at PE diagnosis ≥30 %, no major thrombophilia), we found that patients with RPVO, defined as PVO index ≥5% six months after PE diagnosis, had similar clot formation and clot lysis parameters measured with CLA but higher TGFß1 plasma levels after warfarin discontinuation compared to patients without RPVO. Regarding the risk of recurrent VTE, a shorter clot formation measured with CLA one month after warfarin discontinuation was associated with VTE recurrence during a median follow-up of 41 months. Moreover, time to peak was independently associated with VTE recurrence. In our study, we did not find evidence for an association of plasma fibrinolytic activity, as measured by CLA or assessed by specific profibrinolytic proteins (tPA, uPA, TFPI), with VTE recurrence.

Two previous studies used CLA to assess CLT after a PE episode and analysed the association between CLT and the presence of RPVO. After a first PE episode, Lami *et al.* reported that the presence of RPVO, characterised by a PVO index ≥10% one year after the thrombotic event, was not associated with a longer CLT, which is consistent with our results [21]. Recently, Stepień *et al.* evaluated CLT in RPVO patients, characterised by a PVO index ≥5% three months after a first PE episode, and compared them to patients without RPVO during/after a three-month follow-up. CLT measurements on admission before any anticoagulation and five to seven days following the event were longer in RVPO patients. However, CLT measured three months after the thrombotic event was similar between RPVO and non-RPVO patients [22]. Taken together, these results support the absence of association between RPVO and CLT measured more than three months after PE, in line with the findings of our study. Moreover, no association was found between other clot lysis parameters measured with the CLA (t50% lysis, max lysis rate, time to max lysis rate) and the presence of RPVO.

The association between fibrinolytic biomarkers and RPVO was also investigated. Similarly to our observations, tPA plasma levels were not found to be associated with RPVO, whether they were measured three months or one year after PE [16,21]. In addition, in our study, uPA plasma levels were similar between patients with or without RPVO. The association between uPA plasma levels and the presence of RPVO was not investigated in previous studies focusing on RPVO [16,21,22,33]. TFPI plasma levels were previously described as significantly increased in patients with RPVO [16]. We did not observe a significant difference in TFPI plasma levels between patients with or without RPVO, possibly due to a low number of patients tested.

Regarding clot formation parameters, the association between RVPO and prothrombotic phenotype was previously investigated using turbidimetric clotting assay [21] or calibrated automated thrombography [22]. A shorter lagphase was observed in patients with RPVO compared to patients without [21]. Endogenous thrombin potential was equivalent regardless of RPVO status [22]. While we used a different technique to evaluate clot formation, we found similar clot formation parameters measured with CLA, in particular similar lagphase and peak amplitude, in the RPVO and non-RPVO groups.

Hypofibrinolysis is not the only mechanism that can contribute to RPVO. Obstruction of the pulmonary arteries by unresolved fibrotic clots is described in CTEPH [34]. This accumulation of fibrotic material could be the result of mechanisms involving endothelial cells. Bochenek *et al.* have recently reported that human pulmonary artery endothelial cells overexpressing TGFβ1 acquire a fibroblast-like phenotype [35]. Importantly, when mixed with whole blood, these cells promote the formation of larger clots *in vitro* compared to normal endothelial cells [35]. Moreover, exogenous administration of recombinant TGFβ1 in mice delayed venous thrombus resolution [35]. Significantly elevated TGFβ1 plasma levels were observed in 27 patients with CTEPH compared to 23 patients with pulmonary arterial hypertension and to six healthy controls [35]. Importantly, analysis of plasma levels before and at 12-month follow-up after pulmonary endarterectomy revealed a significant reduction of plasma levels of TGFβ1 [35]. We observed higher TGFβ1 plasma levels and similar fibrinolytic parameters in patients with RPVO compared to patients without RPVO one month after warfarin discontinuation. This could suggest that RPVO might be associated with fibrotic remodeling rather than impaired fibrinolysis. This association between TGFβ1 plasma levels and RPVO seems even stronger in patients presenting recurrent VTE. accordingly, we observed a positive correlation between TGFβ1 plasma levels and RPVO in these patients. Taken all together, these results suggest that TGFβ1 plasma levels could be a dynamic indicator of RPVO.

Global plasma fibrin clot lysis markers such as CLA have been mainly evaluated in order to explore VTE recurrence [11,17]. The most frequently studied CLA parameter is CLT, which is inconsistently associated with VTE recurrence in the literature. In the study of Zabczyk *et al*, including 156 patients with a first episode of PE (provoked or not), prolonged CLT was found to be associated with VTE recurrence [36]. In contrast, in a prospective study of 704 patients with a first unprovoked VTE episode and no thrombophilia, Traby *et al.* reported prolonged CLT as weakly associated with VTE recurrence and only in women [20]. In the study of Meltzer *et al.* performed in 474 patients with a first DVT, there was no association between CLT evaluated three months after discontinuation of oral anticoagulant and the risk of recurrence [37]. Consistent with these two latter studies, we did not observe an association between CLT measured one month after warfarin discontinuation and VTE recurrence after a first unprovoked PE event.

Data concerning clot formation parameters and recurrent VTE are conflicting. Two independent studies used a thrombin-based method to assess lagphase and maximum absorbance of a turbidity curve to explore the potential association between clot formation parameters and VTE recurrence [19,36]. In a population of patients diagnosed with DVT, lagphase was found to be shorter and maximum absorbance to be higher in patients presenting recurrent VTE compared to those without recurrence [19]. There was no difference for lagphase and maximum absorbance regarding recurrence status in a population of PE patients [36]. In contrast, time to peak measured with a calibrated automated thrombogram was shorter in PE patients presenting recurrent VTE, but was similar in DVT patients with or without recurrence. We found a similar procoagulant pattern evaluated with CLA with a shorter lagphase, a shorter time to max CFR, and a shorter time to peak in patients presenting recurrent VTE. In addition, we observed that analysing the most contributing parameters of CLA in PCA identified patients presenting recurrent VTE.

Finally, our findings add to the current knowledge on pathophysiology of RPVO without pulmonary hypertension and of recurrent VTE in patients with unprovoked PE. The observation of an association between TGFβ1 and the presence of RPVO without pulmonary hypertension suggests that elevated TGFβ1 plasma levels could be a marker of early fibrotic remodeling. This fibrotic remodeling might promote recurrent VTE; whether it will promote the development of further CTPEH or not remains to be determined. Therefore, it is plausible that antifibrotic agents might represent a promising adjunctive therapeutic perspective to treat patients with RPVO. For example, nintedanib, a tyrosine kinase inhibitor approved for the treatment of idiopathic pulmonary fibrosis, has been successfully found to reduce pulmonary fibrosis in a mouse model of systemic sclerosis [38]. However, more evidence is needed to support this hypothesis. In addition, the observation of an association between clot formation and lysis parameters and recurrent VTE opens new perspective in order to discriminate more accurately which patients after an unprovoked PE are at high risk of recurrent VTE and which are at low risk. Such progress in the phenotyping of these patients is crucial to determine the optimal duration of anticoagulation.

Our study has however several limitations. First, the sample size was small. Second, we used a tissue factor-based previously published [28,39] rather than a thrombin-based turbidity assay. However, our turbidity assay showed good reproducibility. Some strengths include the careful selection of patients: (i) with a first PE episode (i.e., no previous PE, not proximal DVT) in order to avoid confusion effect of previous RPVO; (ii) that was completely unprovoked (no major transient risk factors, no cancer, no major thrombophilia) to avoid interaction with heterogeneous clinical and biochemical risk factors; (iii) with a significant PVO index at PE diagnosis in order to discriminate between strong responders (no PVO index at 6 months) and partial responders at 6 months (PVO index at 6 months ≥5%). The patients analysed had a strong thrombotic phenotype and were selected from a multicentre double-blind randomised study, with high-quality clinical and biological long-term follow up.

## Conclusion

Among patients with a first episode of unprovoked PE, TGFß1 plasma levels measured one month after VKA discontinuation were found to be associated with RPVO while fibrinolysis parameters were not. Clot formation (lagphase, time to peak absorbance) and clot lysis (maximum lysis rate) parameters measured by CLA were associated with recurrent VTE. These findings, which need to be confirmed in a larger prospective cohort, add to the understanding of the pathophysiology of RPVO and recurrent VTE in order to optimize the treatment of unprovoked PE.

## Data Availability

All data produced in the present study are available upon reasonable request to the authors

## Acknowledgements

The authors would like to thank Dr Julie Larsen from Aarhus University for her help and expert advice during the development of the CLA in our laboratory.

## CRediT authorship contribution statement

**Marc Danguy des Deserts:** Conceptualisation, Methodology, Investigation, Project administration, Resources, Formal analysis, Visualisation, Validation, Writing – original draft, Writing – review and editing. **Claire de Moreuil:** Conceptualisation, Methodology, Validation, Writing – original draft, Writing – review and editing. **Jamal Elhasnaoui:** Formal analysis, Visualisation, Writing – original draft, Writing – review and editing. **Lenaick Gourhant:** Investigation, Resources, Writing – review and editing. **Virginie Gourdou-Latyszenok:** Investigation, Resources, Writing – review and editing. **Benjamin Espinasse:** Writing – review and editing. **Juliette Menguy:** Writing – review and editing. **Cécile Tromeur:** Writing – review and editing. **Rozenn Le Corre:** Writing – review and editing. **Raphael Le Mao:** Writing – review and editing. **Daniel Kraemmer:** Software, Writing – review and editing. **Olivier Sanchez:** Investigation, Writing – review and editing. **Francis Couturaud:** Conceptualisation, Methodology, Supervision, Funding acquisition, Formal analysis, Writing – original draft, Writing – review and editing. **Catherine A. Lemarie:** Conceptualisation, Methodology, Project administration, Funding acquisition, Formal analysis, Validation, Writing – original draft, Writing – review and editing.

## Data availability

Dr Francis Couturaud takes responsibility for data access and integrity of the data. All data produced in the present study are available upon reasonable request to the authors.

## Abbreviations

CFR: clot formation rate
CLA: clot lysis assay
CLT: clot lysis time
CTEPH: chronic thromboembolic pulmonary hypertension
DVT: deep vein thrombosis
GETBO: groupe d’étude de la thrombose en Bretagne occidentale
IQR: interquartile range
OD: optical density
PADIS-PE: prolonged anticoagulation during eighteen months versus placebo after initial six month treatment for a first episode of idiopathic pulmonary embolism
PE: pulmonary embolism
PCA: principal component analysis
PVO: pulmonary vascular obstruction
RPVO: residual pulmonary vascular obstruction
TGFß1: transforming growth factor beta 1
TFPI: tissue factor pathway inhibitor
tPA: tissue plasminogen activator
uPA: urokinase-like plasminogen activator
VKA: vitamin K antagonist
VTE: venous thromboembolic disease
V/Q: ventilation/perfusion

