## Supplementary material for "TGFβ1 plasma levels and clot lysis assay to better characterize patients after a first unprovoked episode of pulmonary embolism"

**Supplementary Figure 1.** Turbidimetry curve and CLA parameters

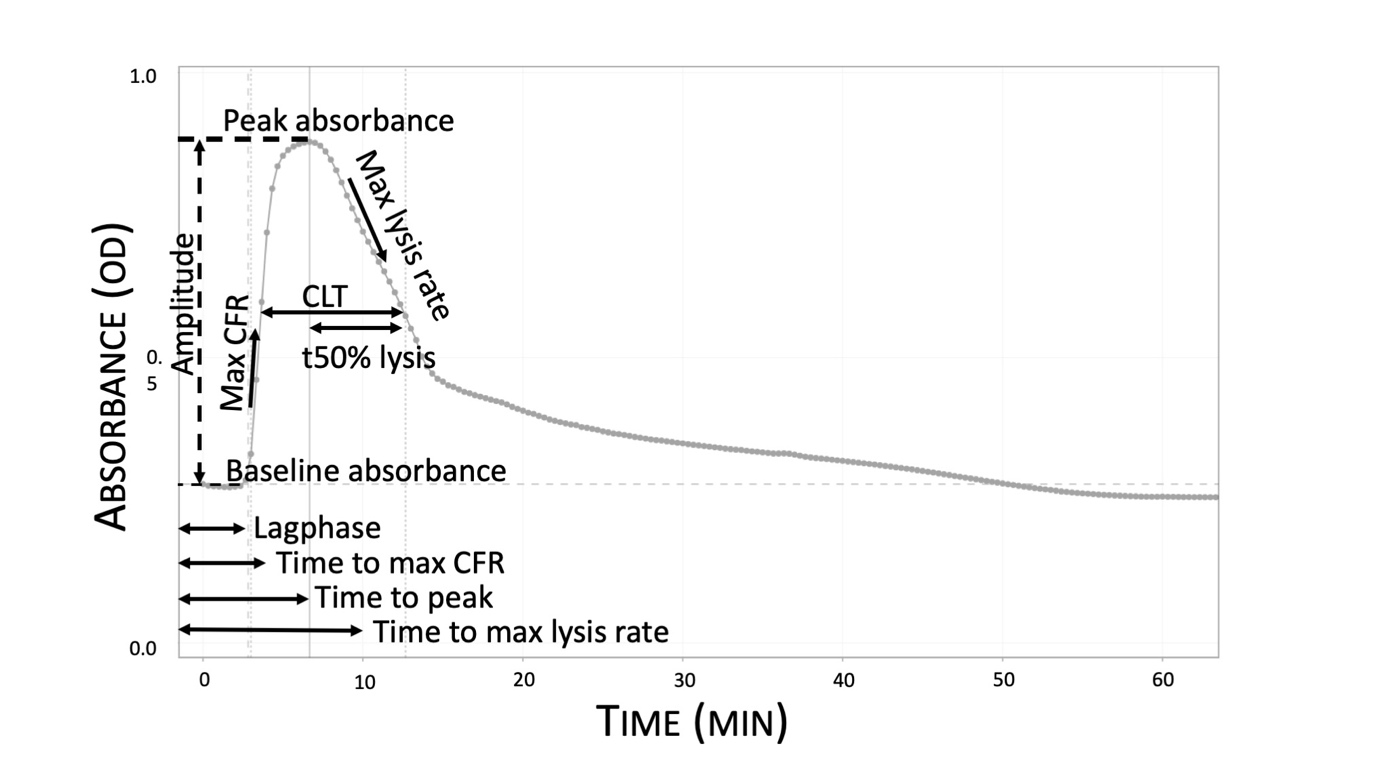

Abbreviations: CFR, Clot formation rate; CLA, Clot lysis assay; CLT, Clot lysis time; OD, Optical density; t50% lysis, time to 50% lysis

**Supplementary Materials and Methods**

**Section 1.** CLA validation and reproducibility study

The aim of this validation study was to evaluate the repeatability and reproducibility of a fibrinolysis assessment technique, the semi-automated Clot and Lysis Assay (CLA), for the measurement of CLT.

Five healthy volunteers were enrolled to form a control plasma pool. 75 µL of frozen plasma were placed in a 96-well plate. An activation solution was prepared as described by Posch (*Transl. Res.* 2020) with a mixture of phospholipids (4 µM), tissue factor (1.375 pM), calcium (20 mM) and tPA (333 ng/mL) in HBSA buffer (HEPES 20 mM, sodium 150 mM, bovine serum albumin 1%, pH 7.4). 75 µL of this solution was added to each well using an OD reader injector (Varioskan Flash, ThermoFisher). The reaction was performed at 37°C, the OD of the wells was measured at a wavelength of 405 nm every 20 seconds for 2 hours. Supplementary Figure 1 illustrates the different parameters calculated from the turbidity curve.

Ten CLA runs were performed. Intra-assay repeatability was assessed using the coefficient of variation (CV in % = standard deviation/mean*100) of the different parameters calculated during the first run of 60 replicas. Inter-assay reproducibility was assessed using the CV of the parameters calculated on the first two replicas of 10 different runs performed by the same operator. Supplementary Table 1 summarizes the obtained CV.

**Supplementary Table 1.** CLA validation and reproducibility results

| **CLA parameter** | **Mean (SD)** | **Intra-assay CV of controls** | **Inter-assay CV of controls** |
| --- | --- | --- | --- |
| Baseline, OD | 0.28 (0.01) | 2.4% | 4.1% |
| Lagphase, sec | 304 (11) | 6.4% | 11.2% |
| Max CFR, OD.min^-1^ | 0.363 (0.025) | 4.7% | 8.0% |
| Time to max CFR, sec | 321 (40) | 6.5% | 12.4% |
| Peak absorbance, OD | 0.87 (0.03) | 2.6% | 3.5% |
| Time to peak absorbance, sec | 521 (29) | 3.7% | 5.5% |
| Amplitude, OD | 0.59 (0.03) | 3.1% | 4.9% |
| Clot lysis time, min | 8.6 (1.0) | 4.6% | 12.1% |
| Time to 50% clot lysis, min | 6.0 (0.9) | 5.9% | 15.9% |
| Max lysis rate, OD.min^-1^ | 0.07 (0.007) | 5.4% | 10.5% |
| Time to max lysis, min | 10.6 (0.7) | 10.4% | 6.6% |

Abbreviations: CFR, Clot formation rate; CLA, Clot lysis assay; CV, Coefficient of variation; OD, Optical density; SD, Standard deviation

**Supplementary Figure 2.** Missing data

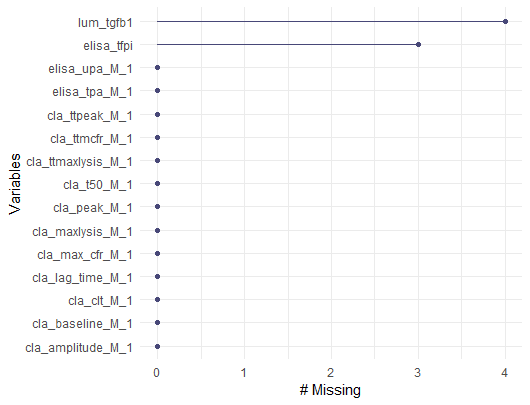

Abbreviations: cfr, clot formation rate; cla, clot lysis assay; clt, clot lysis time; t50, time to 50% lysis, TGFB1, transforming growth factor beta 1; TFPI, tissue factor pathway inhibitor; tPA, tissue-type plasminogen activator ; ttmaxlysis, time to maximum lysis rate, ttmcfr, time to maximum cfr; ttpeak, time to peak; uPA, urokinase-type plasminogen activator;

**Supplementary Table 2.** Group characteristics

|  | RPVO (-) Recurrence (-)  (n=10) | RPVO (-)  Recurrence (+)  (n=7) | RPVO (+)  Recurrence (-)  (n=4) | RPVO (+)  Recurrence (+)  (n=2) | *p-*value |
| --- | --- | --- | --- | --- | --- |
| Age, y | 52.0 (47.5-55.8) | 58.0 (56.0-63.0) | 54.5 (49.3-58.3) | 61.0 (60.5-61.5) | 0.07 |
| Female gender | 4 (40) | 2 (29) | 1 (25) | 0 (0) | 0.71 |
| Body mass index, kg.m^-2^ | 27.9 (25.4-30.6) | 25.8 (24.6-30.0) | 29.3 (26.7-31.3) | 26.6 (25.3-28.0) | 0.92 |
| Creatinine clearance, mL.min^-1#^ | 145 (124-175) | 97 (86-108) | 119 (84-155) | 71 (70-72) | 0.07 |
| Allocated treatment at inclusion |  |  |  |  |  |
| Placebo | 7 (70) | 4 (57) | 3 (75) | 2 (100) | 0.70 |
| Warfarin | 3 (30) | 3 (43) | 1 (25) | 0 (0) |  |
| PVO index at PE diagnosis | 50.0 (47.5-58.8) | 57.5 (52.5-68.8) | 61.5 (60.0-65.4) | 52.5 (41.3-63.8) | 0.23 |
| Biological assessment one month after warfarin withdrawal |  |  |  |  |  |
| Clot Lysis Assay parameters |  |  |  |  |  |
| Baseline, OD | 0.46 (0.33-0.53) | 0.49 (0.40-0.86) | 0.37 (0.33-0.44) | 0.39 (0.37-0.40) | 0.68 |
| Lagphase, sec | 264 (253-283) | 206 (195-235) | 351 (279-400) | 191 (164-217) | 0.07 |
| Max CFR, OD.min^-1^ | 0.31 (0.23-0.37) | 0.32 (0.27-0.44) | 0.27 (0.24-0.31) | 0.35 (0.31-0.39) | 0.65 |
| Time to max CFR, sec | 300 (245-340) | 200 (180-230) | 375 (283-445) | 205 (178-233) | 0.03 |
| Peak absorbance, OD | 1.13 (1.10-1.19) | 1.21 (1.07-1.43) | 1.05 (0.97-1.14) | 0.96 (0.91-1.01) | 0.28 |
| Time to peak absorbance, sec | 525 (493-603) | 460 (435-470) | 590 (505-665) | 395 (373-418) | 0.03 |
| Amplitude, OD | 0.72 (0.64-0.79) | 0.64 (0.52-0.74) | 0.68 (0.63-0.70) | 0.57 (0.51-0.63) | 0.64 |
| Clot Lysis Time, min | 10.7 (10.0-12.3) | 12.3 (11.0-14.2) | 10.1 (9.6-10.8) | 10.5 (10.4-10.6) | 0.22 |
| Time to 50% clot lysis, min | 7.6 (7.0-9.4) | 9.5 (8.1-10.7) | 7.0 (6.3-7.9) | 7.8 (7.6-8.0) | 0.22 |
| Max lysis rate, OD.min^-1^ | 0.07 (0.06-0.08) | 0.05 (0.05-0.06) | 0.07 (0.06-0.09) | 0.05 (0.04-0.06) | 0.08 |
| Time to max lysis rate, min | 15.1 (13.5-16.5) | 15.3 (14.9-17.3) | 15.3 (13.8-17.1) | 9.1 (9.0-9.1) | 0.27 |
| Fibrinolysis markers |  |  |  |  |  |
| D-dimer, ng.mL^-1^ | 647 (408-989) | 463 (221-832) | 293 (199-1108) | 564 (352-765) | 0.81 |
| TFPI, ng.mL^-1^ | 52.3(42.3-60.5) | 46.8 (34.5-56.8) | 48.8 (32.8-53.6) | 53.0 (44.8-61.2) | 0.81 |
| tPA, ng.mL^-1^ | 5.4 (4.4-6.9) | 5.6 (4.3-10.6) | 5.6 (3.8-8.5) | 5.9 (5.9-5.9) | 0.99 |
| uPA, pg.mL^-1^ | 1088 (982-1636) | 1047 (802-1285) | 879 (771-1187) | 725 (684-766) | 0.21 |
| Remodeling marker |  |  |  |  |  |
| TGFß1, pg.mL^-1^ | 66.4 (40.7-78.9) | 89.1 (65.9-116.2) | 97.5 (96.5-121.4) | 225.4 (199.7-251.1) | 0.04 |

Continuous variables are summarized as medians (interquartile range IQR), categorical variables are reported as absolute frequencies (percentages). CFR: clot formation rate; DVT: deep vein thrombosis; OD: optical density; PE: pulmonary embolism; PVO: pulmonary vascular obstruction; TGFB1: transforming growth factor beta 1; TFPI: tissue factor pathway inhibitor; tPA: tissue-type plasminogen activator ; uPA: urokinase-type plasminogen activator; VKA: vitamin K antagonist; VTE: venous thromboembolic disease. ^#^: creatinine clearance was calculated using the Cockroft-Gault formula.

**Supplementary Table 3.** Eigenvalues and cumulative proportion of explained variance by each principal component.

|  | Eigenvalue | Percent explained variance | Cumulative variance percent |
| --- | --- | --- | --- |
| PC1 | 3,93 | 24,54 | 24,54 |
| PC2 | 3,69 | 23,06 | 47,60 |
| PC3 | 2,55 | 15,95 | 63,55 |
| PC4 | 1,80 | 11,27 | 74,82 |
| PC5 | 1,36 | 8,49 | 83,32 |
| PC6 | 0,74 | 4,61 | 87,93 |
| PC7 | 0,62 | 3,84 | 91,77 |
| PC8 | 0,50 | 3,10 | 94,87 |
| PC9 | 0,36 | 2,23 | 97,10 |
| PC10 | 0,23 | 1,41 | 98,51 |
| PC11 | 0,14 | 0,89 | 99,39 |
| PC12 | 0,06 | 0,38 | 99,77 |
| PC13 | 0,03 | 0,17 | 99,94 |
| PC14 | 0,01 | 0,06 | 100,00 |
| PC15 | 0,00 | 0,00 | 100,00 |
| PC16 | 0,00 | 0,00 | 100,00 |

The first five principal components with eigenvalues > 1 were retained for further analysis.

**Supplementary Figure 3.** Contribution of variables to PC1 and/or PC2.

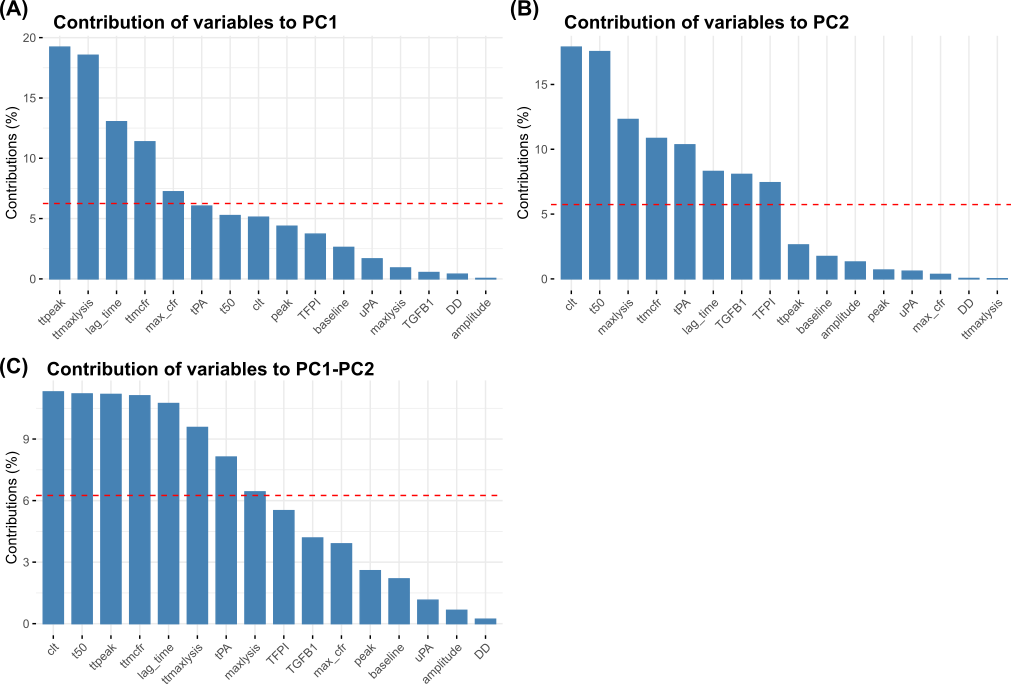

The contribution (in percentage) of each of the 16 parameters to (A) the first, (B) the second, and (C) to both principal components. The red dashed lines represent average expected contribution. If the contribution of the variables is uniform, the expected average contribution value should be 1/16 = 6.25%. The variables with a contribution greater than this value are important contributors to the component.
